# Detection of maternal transmission of resistant Gram-negative bacteria in a Cambodian hospital setting

**DOI:** 10.1101/2022.10.07.22280813

**Authors:** Chalita Chomkatekaew, Janjira Thaipadungpanit, Pasco Hearn, Sona Soeng, Sreymom Pol, Leakhena Neou, Jill Hopkins, Paul Turner, Elizabeth M. Batty

## Abstract

Infection with Extended-spectrum beta-lactamase -producing *Enterobacterales* (ESBL-E) is common in infants leading to increased intensive care unit admission and mortality, but the role of maternal transmission in colonisation of infants is unclear. Using paired isolates from 50 pairs of mothers and neonates admitted to a Cambodian hospital, we investigated antimicrobial resistance in *Escherichia coli* and *Klebsiella pneumoniae* using whole genome sequencing. We detected a wide variety of ESBL-E genes present in this population along with high levels of multidrug resistance. From 21 pairs where the same organism was present in both mother and neonate, we identified eight pairs with identical or near-identical isolates from both individuals suggestive of transmission at or around birth, including a pair with transmission of multiple strains. We found no evidence for transmission of plasmids only from mother to infant. This suggests vertical transmission outside hospitals as a common cause of ESBL-E colonisation in neonates.

## Introduction

Antimicrobial resistance (AMR) is a major global health problem, and multiple studies have shown high rates of AMR in low and middle-income countries in Southeast Asia compared to other regions [1–3]. AMR is a particular problem in neonates, where infection with Extended-spectrum beta-lactamase -producing *Enterobacterales* (ESBL-E) are common and associated with admission to ICU and death [4–6]. ESBL-E infections have become one of the main challenges for antibiotic treatment leading to the increase in mortality, healthcare cost and threatening the effectiveness of first line sepsis treatments in low resource countries such as Cambodia. Rates of ESBL-E in Cambodia have been shown to rise between 2012 and 2015. However, multidrug resistance infection cases in Cambodia may be under-reported compared to neighboring countries due to lack of infrastructure [7], and AMR data in Cambodia is improving but currently limited [1].

Cambodia shows a high prevalence of ESBL-E colonisation in the early neonatal period [8] suggestive of mother-to-child transmission at or shortly after delivery. A high number of infants are likely to already be colonised prior to hospital admission, and community acquisition may be important in the neonatal population [9]. Maternal transmission has been implicated in ESBL-E acquisition by neonates by looking at prevalence [10], or by using matched antimicrobial resistance phenotypes [11], pulsed-field gel electrophoresis [12], repetitive element PCR typing [13] to compare strains between mother and infant. Most recently, the BARNARDS study identified identical isolates between mother and infant in *E. coli* using whole-genome sequencing, suggesting mother-to-child transmission in a large south Asian and African dataset [14].

Whole-genome sequencing (WGS) of clinical isolates has emerged as an invaluable tool with many applications in clinical microbiology including tracking outbreaks, monitoring trends in infections and investigating pathogen transmission routes [15]. Several studies have shown that high-resolution data from WGS is a proven tool to track the possible transmission of *Klebsiella pneumoniae* circulating both in the intensive care unit [16] and in an outbreak via plasmid transmission [17]. However, most current datasets are short-read data, and the assembly of short-reads datasets usually results in multiple contigs and ambiguous alignments instead of a complete genome [18]. To overcome this challenge, Oxford Nanopore MinION platform can produce long reads enabling improved quality genome assembly including complete assembly of plasmids [19].

We used whole-genome sequencing to obtain ESBL-E genomes from pairs of mothers and neonates admitted to Angkor Hospital for Children, Siem Reap, Cambodia. Using complete genomes generated using short- and long-read sequencing, we identified patterns of antimicrobial resistance and investigated potential transmission events between mothers and neonates to better understand the importance of maternal transmission versus community acquisition in early colonisation of neonates.

## Methods

### Study location

Angkor Hospital for Children is a non-governmental paediatric referral hospital with a dedicated neonatal intensive care unit (NICU) and special care baby unit (SCBU). The hospital has no maternity unit, thus all neonatal admissions are outborn. Over a one year period all infants aged ≤28 days were eligible for study enrolment on admission to the NICU or SCBU, excluding those who had any previous healthcare service exposure following delivery (e.g. overnight admission to a hospital / health centre or transfer from another AHC ward).

### Sample collection, isolation, and phenotyping

A rectal swab was taken from the infant within 24 hours of admission to the neonatal unit (at a median of 11 days of age [range 0 – 30]), and a maternal stool sample was collected as soon as possible after infant admission. Samples were cultured onsite on selective chromogenic ESBL detection agar (CHROMagar ESBL medium; CHROMagar, France) and all morphotypes followed up for analysis. Species identification was confirmed by standard biochemical tests and antimicrobial susceptibilities were determined by disk diffusion, following CLSI methodology and using 2017 breakpoints [20]. Double disk diffusion tests were done to confirm ESBL production (cefotaxime and ceftazidime with/without clavulanate; BBL, Becton Dickinson, USA).

### Bacterial growth and DNA extraction

96 isolates of *E. coli* and *K. pneumoniae* were cultured from frozen stocks (−80°C) and grown in LB broth at 37°C for 3 hours. 900 μl of bacterial culture was spun down at 21,200 rcf for 5 minutes before the supernatant was removed. The bacterial cells were lysed with 600 μl of Nucleic lysis solution (Promega, USA) at 80°C for 10 minutes. The cell lysate was then transferred to a new tube containing 250 μl of glass beads with 5 μl of RNase A solution (Qiagen, Germany) and the cells were further lysed by vortexing for 10 minutes. The cell lysate was incubated at 37°C for 30 minutes to remove the RNA. The mixture was purified using magnetic beads (Agencourt AMPure XP beads, Beckman Coulter, USA) with a 1:4 volume ratio of lysate mixture and magnetic beads. The extracted DNA was redissolved in distilled sterilised water and then stored in the freezer (−20°C) prior to library preparation for sequencing.

### Sequencing

Sequencing libraries were prepared from extracted DNA using Nextera Flex library preparation kits and sequenced on an Illumina MiSeq with 300bp paired-end reads. 36 isolates were selected for long-read sequencing. The Rapid Barcoding Kit (Nanopore, UK) was used to prepare DNA libraries according to the Nanopore protocol with 12 barcoded samples per flow cell. The prepared libraries were pooled and purified using an additional step with magnetic beads (Agencourt AMPure XP beads, Beckman Coulter, USA) using a 1:1 volume ratio of pooled libraries and the magnetic beads. The purified libraries were then sequenced on a MinION R9.4 flow cell according to the Nanopore protocol. The sequences were deposited in the European Nucleotide Archive (ENA) at EMBL-EBI under project accession number PRJEB37551 https://www.ebi.ac.uk/ena/browser/view/PRJEB37551.

### Bioinformatic analysis

Raw sequence data was processed using the Bactopia pipeline 1.4 [21]. Reads were mapped against *E. coli* K-12 substrain MG1655 reference sequence for all *Escherichia* isolates (https://www.ncbi.nlm.nih.gov/assembly/GCF_000005845.2/) and *K. pneumoniae* HS11286 (https://www.ncbi.nlm.nih.gov/assembly/GCF_000240185.1/) for all *Klebsiella* isolates, and the genome assemblies were also used as references for confirmation of transmission. Mashtree 1.1.2 [22] was used to generate phylogenetic trees from all samples and visualised using ggtree 3.1.2 [23]. Ska 1.0 [24] was used to perform split k-mer analysis using the default cutoffs (95% kmer identity and <20 SNPs to call transmission clusters). Snippy 4.6.0 [25] was used to produce variant calls, and snippy-core was used to mask repetitive regions and produce a core SNP alignment. SNP distances between samples were called with snp-dists 0.6.3 [26]. MLST was determined using mlst 2.19.0 [27] and the PubMLST database [28]. Genome assemblies were performed using Unicycler 0.4.8 [29]. *Escherichia* species and phylogroups were determined using ClermonTyping [30]. *Klebsiella* phylogroups and virulence genes were obtained through Kleborate [31]. Antimicrobial resistance genes and mutations were determined using NCBI AMRFinder+ 3.8.4 [32] and upset plots were visualized using ComplexUpset. Platon 1.5.0 [33] was used to identify contigs originating from plasmids and the mobilization proteins, replication proteins, and incompatibility groups on each contig. The Platon identification was used to determine whether the AMR genes were part of the chromosome or present on plasmids. Flanker 0.1.5 [34] was used to extract 20kb genomic regions surrounding AMR genes, treating plasmids as circular if the Unicycler assembler reported them as circular. Isolates were removed if the AMR gene was on a contig less than 5kb in length. Clinker [35] was used to visualise the regions, with links drawn between genes with 95% or greater identity.

## Results

Between 23rd March 2016 and 13th March 2017, 121 isolates were cultured from swabs from 50 pairs of mothers and infants. 25/50 (50%) of infants and 47/50 (94%) of mothers were colonised with an ESBL-E, including 17/50 pairs where both mother and infant were colonised with *E. coli* and 4/50 pairs colonised with *K. pneumoniae*. Nine pairs had the same species isolated from both mother and infant with identical antibiograms, which we considered as potential transmission pairs (Figure 1).

**Figure 1:**
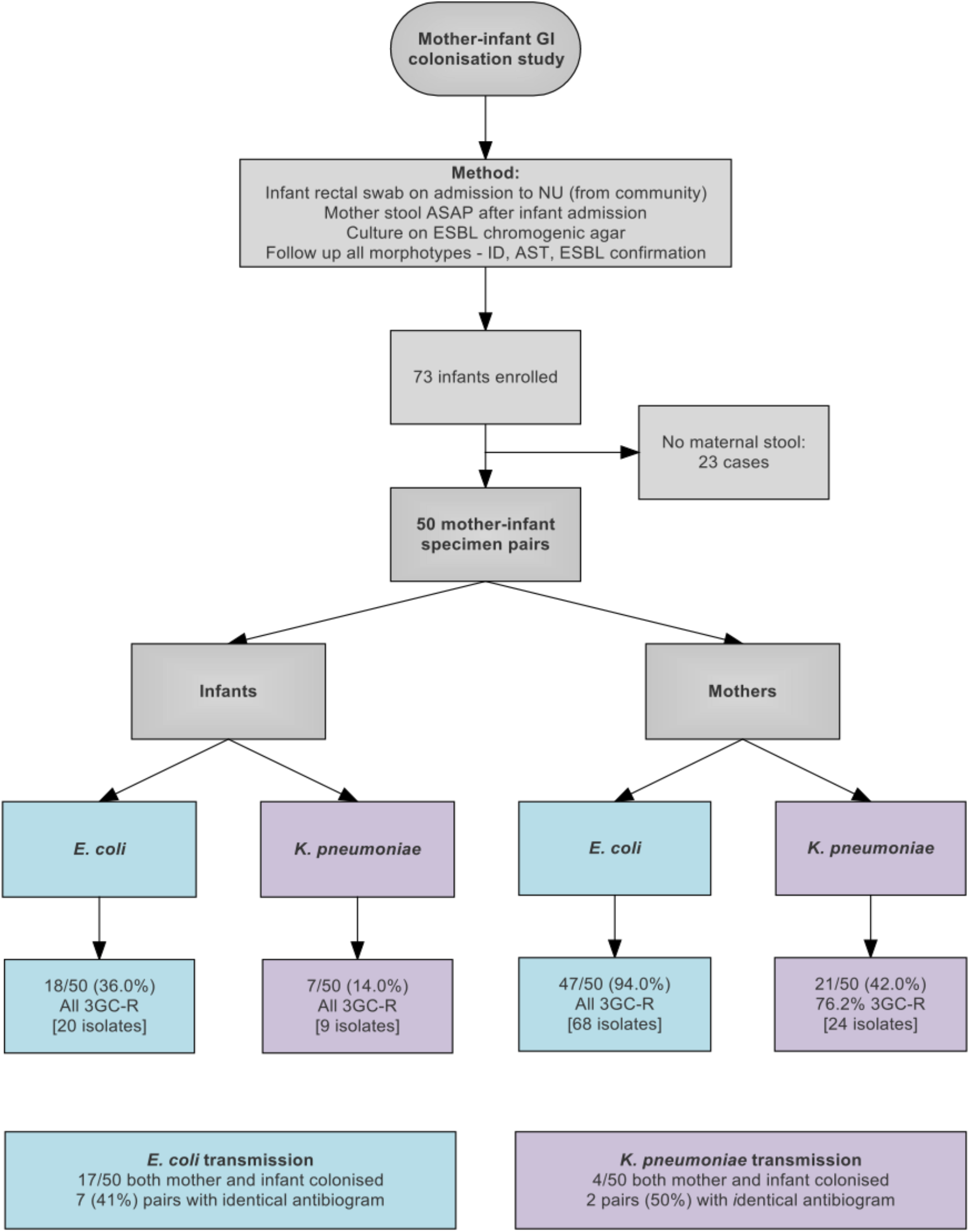
Flow diagram showing study design and sample collection. 3GC-R: third generation cephalosporin resistant.

We sequenced 96 isolates in total on Illumina MiSeq - 68 isolates from 20 of the 21 pairs where both mother and infant were colonised, including eight of nine pairs considered as potential transmission pairs (one pair could not be retrieved for sequencing), and 28 other randomly selected isolates from the study. We sequenced up to four isolates per individual based on the number of distinct morphotypes identified during bacterial growth (Supplementary Table 1).

We performed long-read sequencing on the 18 isolates identified as possible transmission pairs based on the phenotypic data, as well as 18 other isolates which were observed to have closely related core genomes or similar patterns of antimicrobial resistance genes based on Illumina data. We used hybrid assembly of the long and short-read data to obtain complete genomes and resolve the plasmids in these strains in order to confirm transmission events and identify whether these transmission events involved complete isolates, or plasmid horizontal transmission between different isolates. We obtained between 1 and 9 contigs for each of the 36 isolates, including one contig of 4.5Mb or larger assumed to be the chromosome, and 0-8 plasmid contigs per isolate.

The species of each isolate was determined from the whole genome sequencing using ClermonTyping (for *Escherichia* species) and Kleborate (for *Klebsiella species)*. This showed that the *K. pneumoniae* isolates included three isolates of *Klebsiella quasipneumoniae subsp. similipneumoniae* and one isolate of *Klebsiella quasipneumoniae subsp. quasipneumoniae*, and the *E. coli* isolates included three isolates of *Escherichia fergusonii* and two isolates of *Escherichia clade-1* (Figure 2).

**Figure 2:**
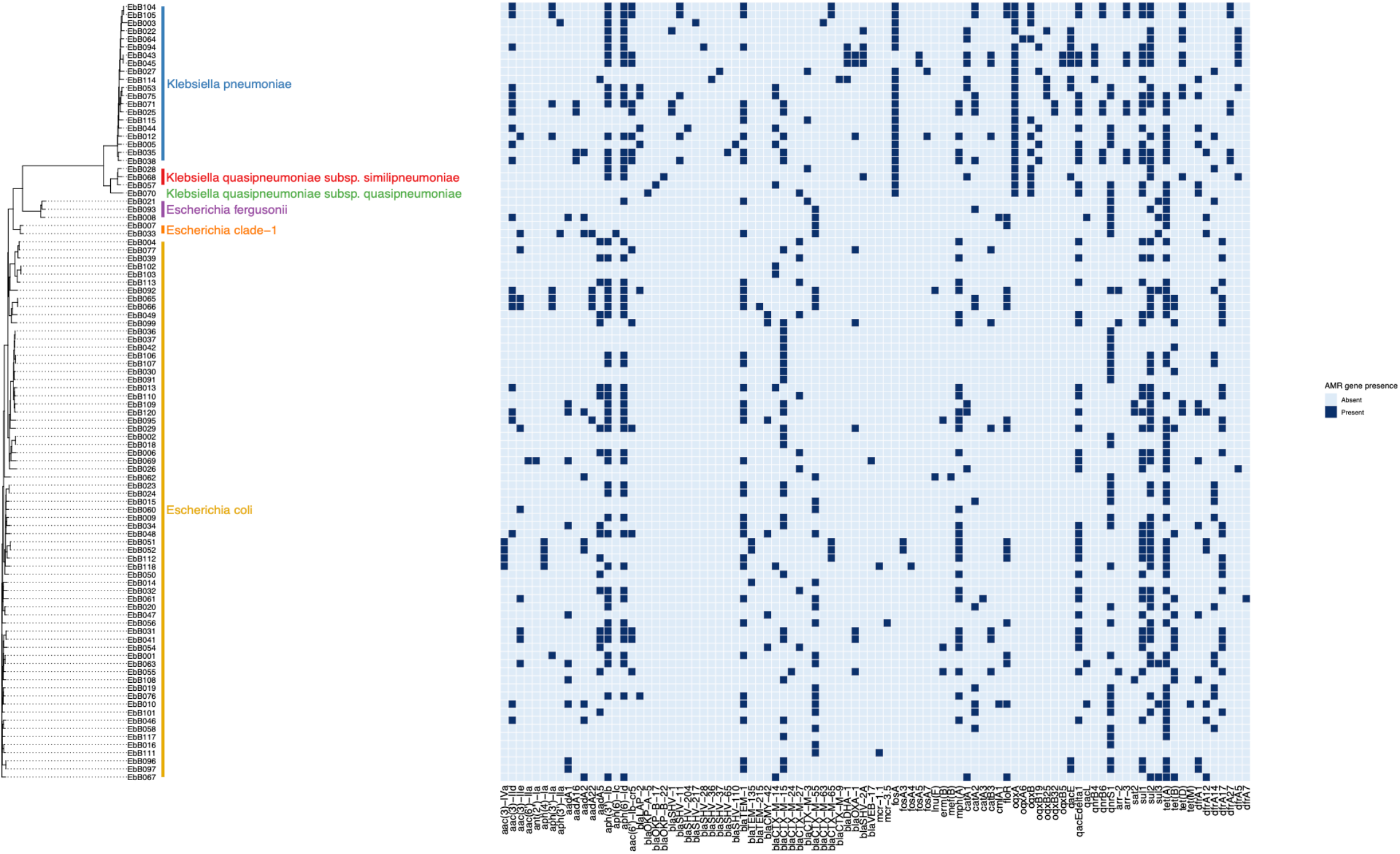
phylogeny based on mashtree of all samples showing species, and presence of selected AMR genes in each sample.

No phylogroups or MLST were predominant in the isolates. Among *E. coli* isolates, the most common MLST was ST38 and the closely related ST3268 and ST3052 and ST318, as well as three isolates from the ST131 pandemic clade (Supplementary Figure 1). Of the three ST131 isolates, two were clade C and one was clade A. In the *K. pneumoniae* isolates there were no dominant sequence types, with ST17 the only sequence type present more than once in unpaired isolates, and four isolates had novel sequence types (Supplementary Figure 2). No virulence determinants were found in the *K. pneumoniae* isolates and all had a virulence score of 0 as determined by Kleborate.

### Antimicrobial resistance

95 of 96 isolates had an ESBL gene present in the genome, as expected as they were selected for the presence of ESBLs. One isolate (EbB064) had no ESBL genes present in the sequenced genome, despite having phenotypic resistance to 3rd generation cephalosporins. Four *Klebsiella pneumoniae* and one *Klebsiella quasipneumoniae subsp. similipneumoniae* isolates had *bla*_SHV-2A_ genes predicted to cause ESBL resistance, with two cases of *bla*_SHV-2A_ carried on a plasmid, one case where the gene is chromosomal, and one pair of isolates carrying the gene on both a plasmid and in the chromosome. Multidrug resistance was extremely common in these isolates, with 64% (61/96) of isolates genotypically resistant to 6 antimicrobial classes, including 2 isolates resistant to 8 classes (Figure 3). No carbapenemase resistance was found in this dataset. Colistin resistance was observed through presence of *mcr-1*.*1* and *mcr-3*.*5* genes in three *E. coli* isolates. A point mutation of *pmrB* (R256G) was reported in three *K. pneumoniae* isolates but this has been previously demonstrated not to cause colistin resistance in isolation [36] and we did not consider these isolates resistant. AMRfinder analysis reported resistance to phenicol/quinolone via *oqxAB* and resistance to fosfomycin via *fosA* in *K. pneumoniae*, but Kleborate confirmed these were intrinsic and no acquired resistance was reported, and so these isolates were not recorded as resistant through presence of these genes [31].

**Figure 3:**
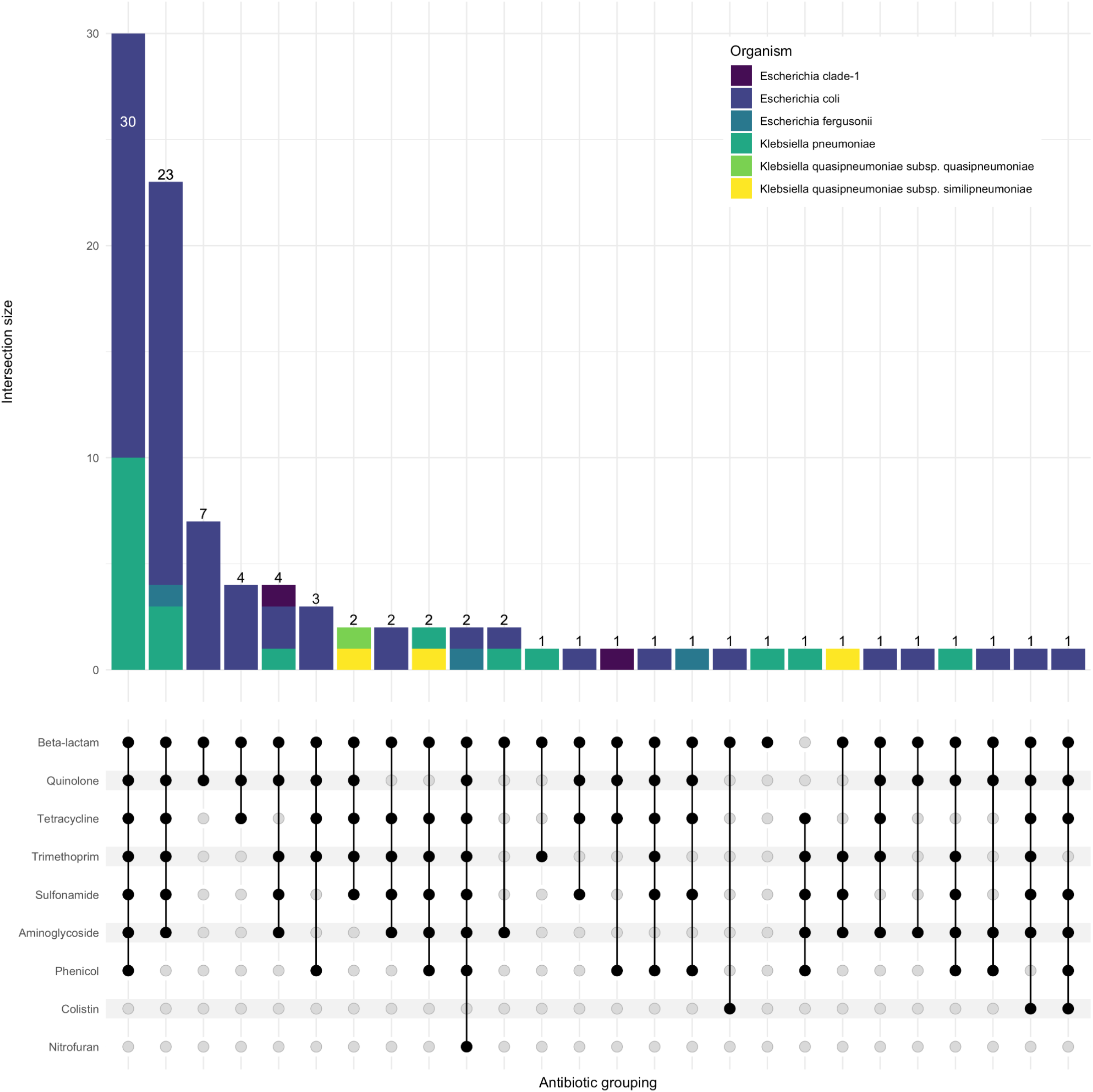
Upset plot showing shared classes of antimicrobial resistance genes between isolates. Isolate counts are shown as bars, with antimicrobial resistance gene combinations displayed in the lower panel.

A wide range of beta-lactamase genes were carried, including 9 different *bla*_CTX-M_ genes, and genes from the *bla*_CMY_, *bla*_OXA_, *bla*_DHA_, *bla*_SHV_ and *bla*_VEB_ families (Figure 4). The most common were *bla*_CTX-M-15_, found in 31 isolates, and *bla*_CTX-M-55_, found in 24 isolates. 25 isolates had an ESBL gene in the chromosome, while 7 isolates had ESBL genes present on plasmids as well as integrated into the chromosome.

**Figure 4:**
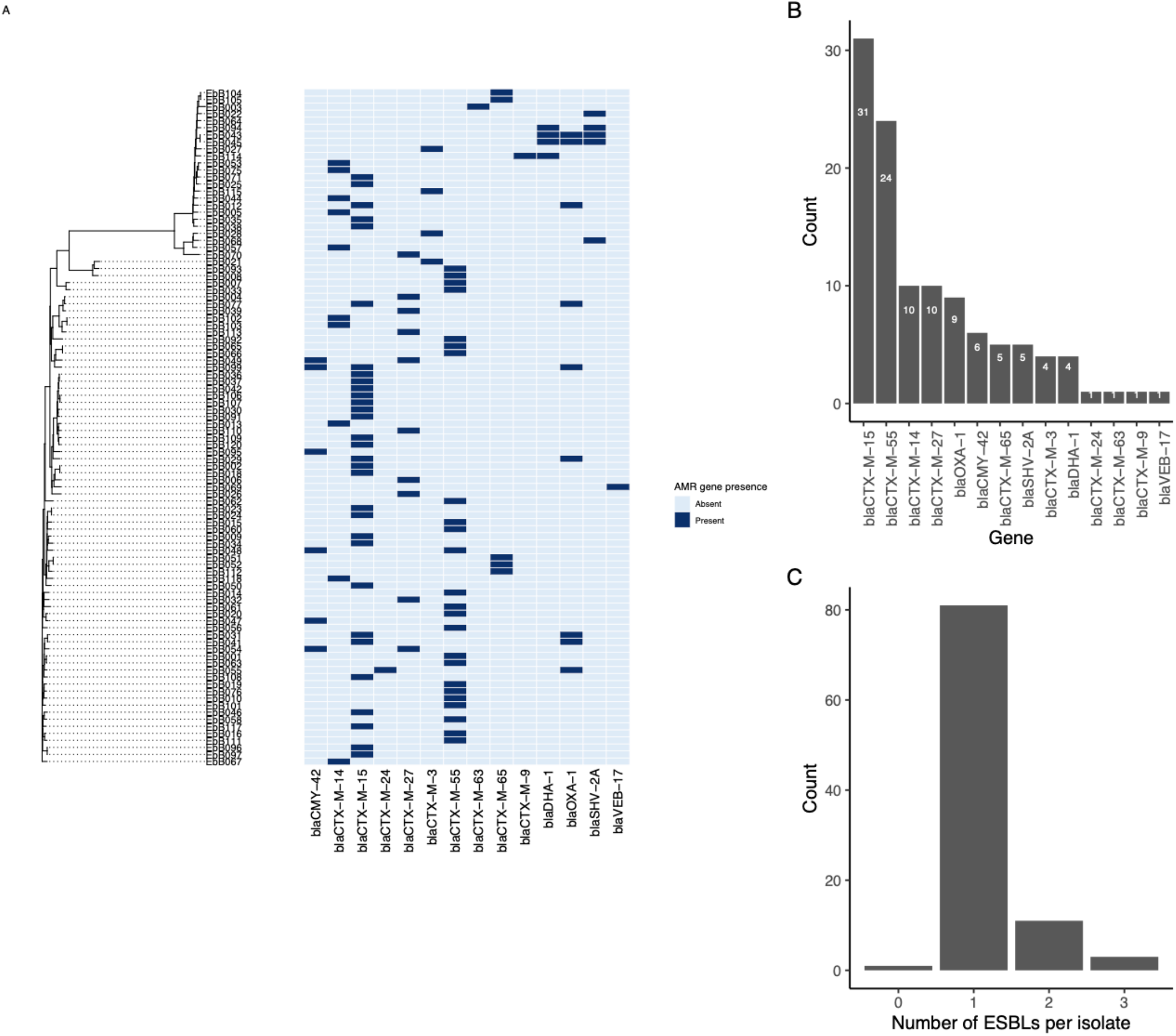
a) Phylogeny from mashtree showing presence/absence of ESBL genes b) frequency of ESBL genes in the isolates and c) number of ESBL genes per isolate.

### Identification of transmission events

For each potential pair of isolates from a mother-infant pair, we identified potential transmission events as either transmission of a strain with complete genome (chromosome only, or chromosome and plasmid), transmission of plasmids, or no transmission.

For eight of the nine pairs which had the same antibiogram in both strains, the mother and infant strains shared the same MLST and the same set of acquired antimicrobial resistance genes, and a further pair of isolates differed only by one antimicrobial resistance gene. We used split k-mer analysis to identify transmission clusters in the complete dataset, and confirmed this using the complete genome assembled from one of the paired strains as a reference for mapping and variant calling. We removed variant calls in repetitive regions but did not correct for recombination as we assume no recombination will occur in the timeframe of maternal transmission.

Eight of these nine pairs were identified as transmission clusters using ska with default filters (95% kmer similarity and a SNP distance threshold of 20), and had 0-4 SNP differences between them (Figure 5). To confirm these results, we used one strain from each pair as a reference genome, and performed mapping and variant calling. Mapping to one pair as a reference gave similar results in all cases with 0-4 SNP differences from the mapping-based approach. We identified one pair where complete strains of both *E. coli* and *K. pneumoniae* were transmitted.

**Figure 5:**
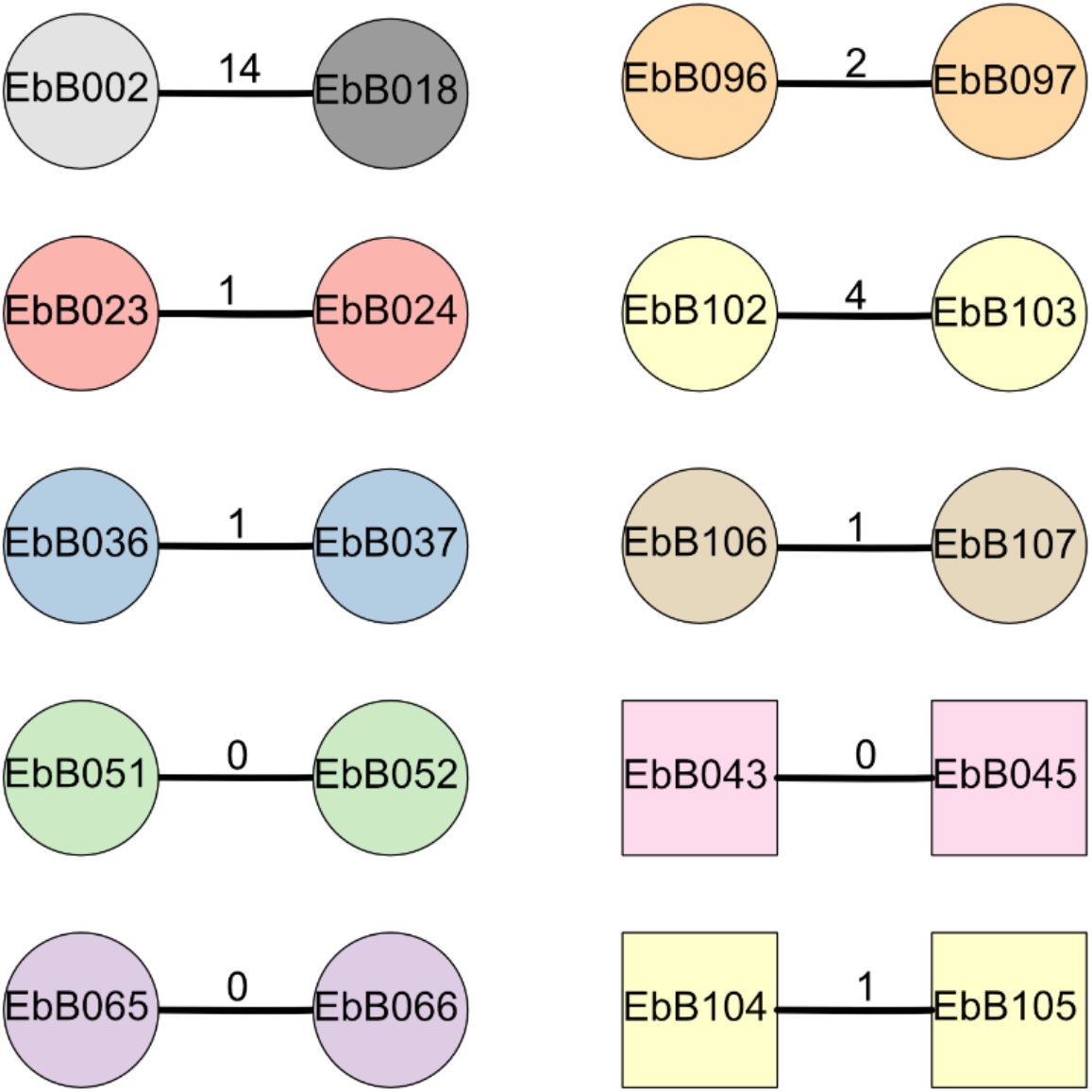
Diagram of clusters identified using ska. Circles show strains of *E. coli* while squares are *K. pneumoniae*. The number on the bar joining each pair represents the SNP distance between the two strains. Shapes are coloured according to the mother-infant pairs they were isolated from, with grey shapes for strains taken from unpaired individuals.

We also looked at two other pairs of isolates from the whole collection which were identified as a possible transmission cluster by ska (Figure 5). In one case, the two isolates (EbB096 and EbB097) were from the same individual, and likely represent the same isolate which was sequenced twice due to appearing as different morphotypes during bacterial growth. The other pair of isolates (EbB002 and EbB018) were identified as a transmission cluster by ska, but with 14 SNP differences between the isolates. Mapping to one of the paired isolates as a reference gave similar results, with 13 SNP differences between isolates. This pair of isolates came from an unpaired mother and infant, and were collected 54 days apart.

For each of the transmission pairs we performed whole genome alignment using minimap to determine if the transmission included plasmids. Seven of the eight transmission clusters included plasmids, with only one pair where only the chromosome was present in both assemblies. While the plasmids appear to be transmitted in all strains, in three of the eight transmitted pairs there were rearrangements between plasmids. In one pair (isolates EbB065 and EbB066), one isolate had two circular contigs of 280kb and 95kb, while the other isolate has only one non-circular contig of 372kb, which likely represents a misassembly of two plasmids into one contig (Supplementary Figure 3). In another pair (EbB043 and EbB045), an 88kb non-circular contig from one isolate is present in the chromosome of the paired isolate, representing either a misassembly, or potentially insertion into the chromosome (Supplementary Figure 4). In a third pair (EbB104 and EbB105), the same plasmid material is assembled into a single plasmid in both strains, but with rearrangements between the plasmids, including a tandem duplication of the *bla*_CTX-M-65_ gene seen in two copies in EbB104 and three copies in EbB105 (Supplementary Figure 5).

We also considered the possibility that maternal transmission of plasmids could occur in the absence of transmission of a complete strain, which would not be discovered using mapping-based approaches but may be identified by similar antimicrobial resistance profiles. To check for plasmid-borne resistance genes shared between strains, we compared the antimicrobial resistance profiles of all strains. There were no cases of identical or near-identical antimicrobial resistance profiles in paired strains which do not have similar core genomes. We did identify three clusters of unpaired samples with identical or near-identical antimicrobial resistance profiles which do not have similar core genomes: EbB004 and EbB039, EbB031 and EbB041, and EbB036/EbB037 and EbB091.

Strains EbB004 has a 127kb plasmid carrying incFII and incFIB, while EbB039 has a 221kb plasmid with incFII, incFIB, col156 and P0111 replicons. The two plasmids shared a 50kb segment with an internal inversion, including the *bla*_CTX-M-27_ gene, and eight other genes conferring resistance to aminoglycosides, trimethoprim, macrolides, sulfonamides, and tetracycline.

Strains EbB031 and Eb041 both belong to ST44 and have a SNP distance of 349 measured by ska. They also share a 170kb plasmid with a similar resistance cassette carrying both *bla*_CTX-M-15_ and *bla*_OXA-1_, as well as resistance to aminoglycosides, trimethoprim, macrolides, sulfonamides, tetracycline, and chloramphenicol.

Strains EbB036 and EbB037 are a transmission pair which share an antimicrobial resistance profile with strain EbB091. However, none of these strains contain plasmids, and the shared resistance profiles are driven by chromosomal *qnrS1* and *bla*_CTX-M-15_ integrated in different locations.

Finally we looked at the genomic context of ESBLs found in more than one strain to determine if shared mobile elements were driving the spread of ESBLs in this dataset. Observing the 20kb regions around each ESBL, many of the ESBL genes were present in different contexts and were not present on shared plasmids or as shared mobile elements integrated into plasmids or chromosomes.

## Discussion

We use complete genomes combining short and long-read sequencing to show that maternal transmission is an important source of neonatal colonisation. We document seven cases of maternal transmission across 50 pairs of mothers and infants, which may be an underestimate if the colony selection on the basis of differing morphology did not represent the full diversity of *E. coli* and *K. pneumoniae* carriage. In eight of nine transmitted strains, the strain was transmitted with all plasmids intact.

We identified potential transmission pairs in this study by looking for similar patterns of phenotypic antimicrobial resistance. Using similar antimicrobial resistance phenotypes in the same organism as the sole criteria for detecting transmission, we would have detected all of the transmission pairs, and called only one false positive transmission. Using similar antimicrobial resistance and requiring the same MLST would have successfully detected all transmission pairs confirmed using complete genomes, and would have excluded the false positive. This suggests that previous studies of maternal transmission using MLST or PFGE genotyping to identify transmissions are likely to have correctly called transmission events. However, we detected at least one pair of isolates which were closely related but are from individuals from unrelated pairs and do not represent maternal transmission between these individuals. As the samples were collected on admission to the hospital, this is more likely to be community transmission than due to a common source in the hospital. This suggests that community transmission of isolates can give rise to results which are similar to maternal transmission and may confound transmission studies, particularly if a SNP threshold for transmission cannot be easily identified.

The isolates in this study were pre-selected to include only those with ESBLs and may not represent a true population survey. Previous estimates of carriage rates in Cambodia have ranged from 20% in rural villages in 2011 [37], 42.7% for *E. coli* and 33.7% for *K. pneumoniae* between 2012 and 2015 [38], and 92.8% for *E. coli* and 44.1% for *K. pneumoniae* in Siem Reap province in 2019 [39]. Despite the selection for ESBL carriage, the results are largely concordant with previous studies investigating all isolates, with high levels of genetic diversity, no dominant sequence types, and a range of antimicrobial resistance genes [40]. Despite the presence of ST131 isolates in this dataset, they were not dominant in carriage. A wide range of ESBL genes were seen in these isolates, with *bla*_CTX-M-15_ and *bla*_CTX-M-55_ the most common, but no dominant gene or lineage. Previous studies have reported hypervirulent *K. pneumoniae* clones as a cause of community-acquired infection in southeast Asia [41], but we did not detect any *K. pneumoniae* virulence genes in this dataset.

We were unable to find any examples of transmission of plasmids between mothers and infants outside the transmission of a complete strain. Across the whole dataset there were few shared plasmids outside transmission pairs, indicating that no dominant plasmids are responsible for the spread of antimicrobial resistance. The few strains with similar plasmids from unrelated individuals, such as EbB031 and EbB041, were also closely related in the core genome and suggests community circulation of strains rather than plasmid transmission. The complete genome sequences allowed us to look at the different antimicrobial resistance genes in context, and show that the ESBL genes are present in different contexts in both plasmid and chromosome, and that no common transposable element is responsible for antimicrobial resistance transmission in this dataset. Our limited set of ST131 genomes showed the characteristic incF plasmids carrying *bla*_CTX-M-27_, as previously reported as part of the diverse plasmidome of ST131 [42].

Our study suggests that vertical transmission outside the hospital is a common source of neonatal colonisation. Strategies to reduce neonatal infections need to consider that maternal transmission is a frequent occurrence, and that AMR strains are circulating in the community and widely carried by mothers and infants. Strategies which focus solely on hospital-based intervention may be unsuccessful at preventing transmission and colonisation which occurs in the community.

## Data Availability

All sequencing data produced is online at European Nucleotide Archive (ENA) under project accession number PRJEB37551.

https://www.ebi.ac.uk/ena/browser/view/PRJEB37551

## Ethics statement

The study was approved by Angkor Hospital for Children (0034/16-AHC), the Cambodia National Ethics Committee for Health Research (464-NECHR) and the Oxford Tropical Research Ethics Committee (575-15).

## Data summary

Raw sequence data generated in this study were deposited in the European Nucleotide Archive (ENA) at EMBL-EBI under project accession number PRJEB37551 https://www.ebi.ac.uk/ena/browser/view/PRJEB37551

