## Supplementary Table and Figures for "Detection of maternal transmission of resistant Gram-negative bacteria in a Cambodian hospital setting"

### Supplementary Figures

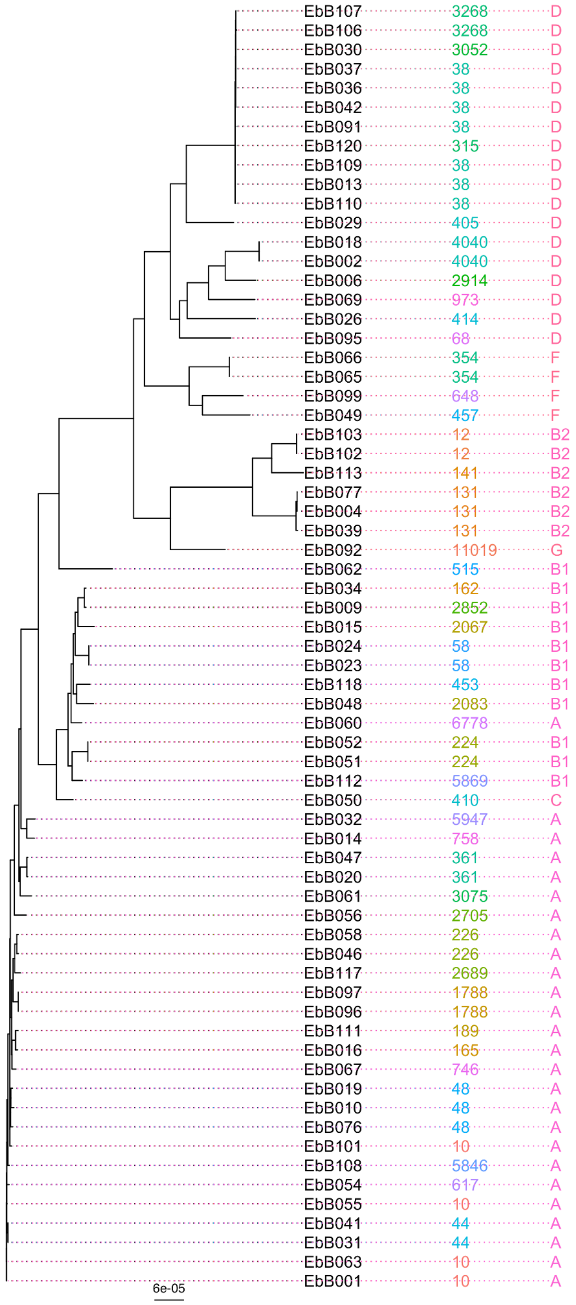

Supplementary Figure 1. A phylogeny of the *E. coli* isolates sequenced in this study including their ST and phylogroup. Variant calls from Snippy were filtered to remove repeat regions and recombinant regions using Gubbins, and the core SNPs were used to generate a tree using IQ-TREE.

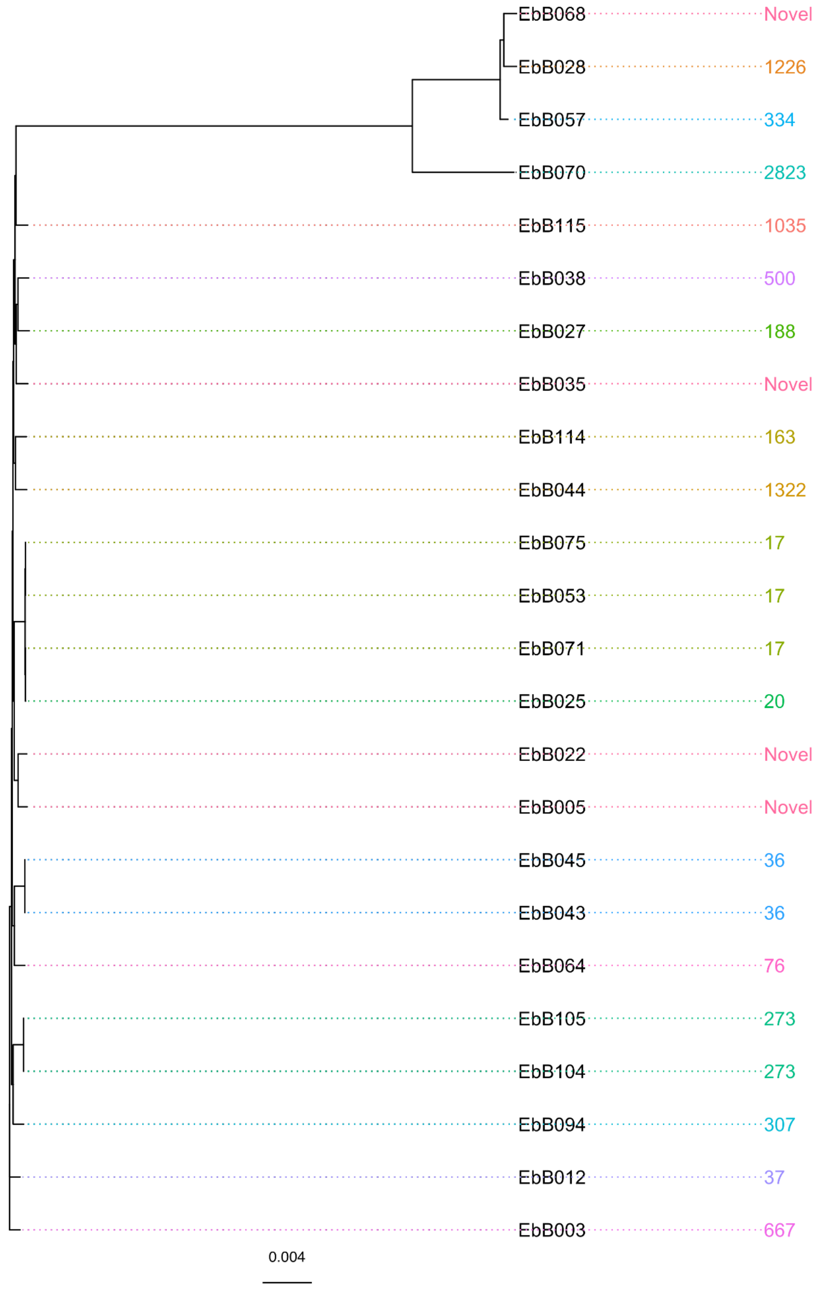

Supplementary Figure 2. A phylogeny of the *K. pneumoniae* isolates sequenced in this study including their ST. Variant calls from Snippy were filtered to remove repeat regions and recombinant regions using Gubbins, and the core SNPs were used to generate a tree using IQ-TREE.

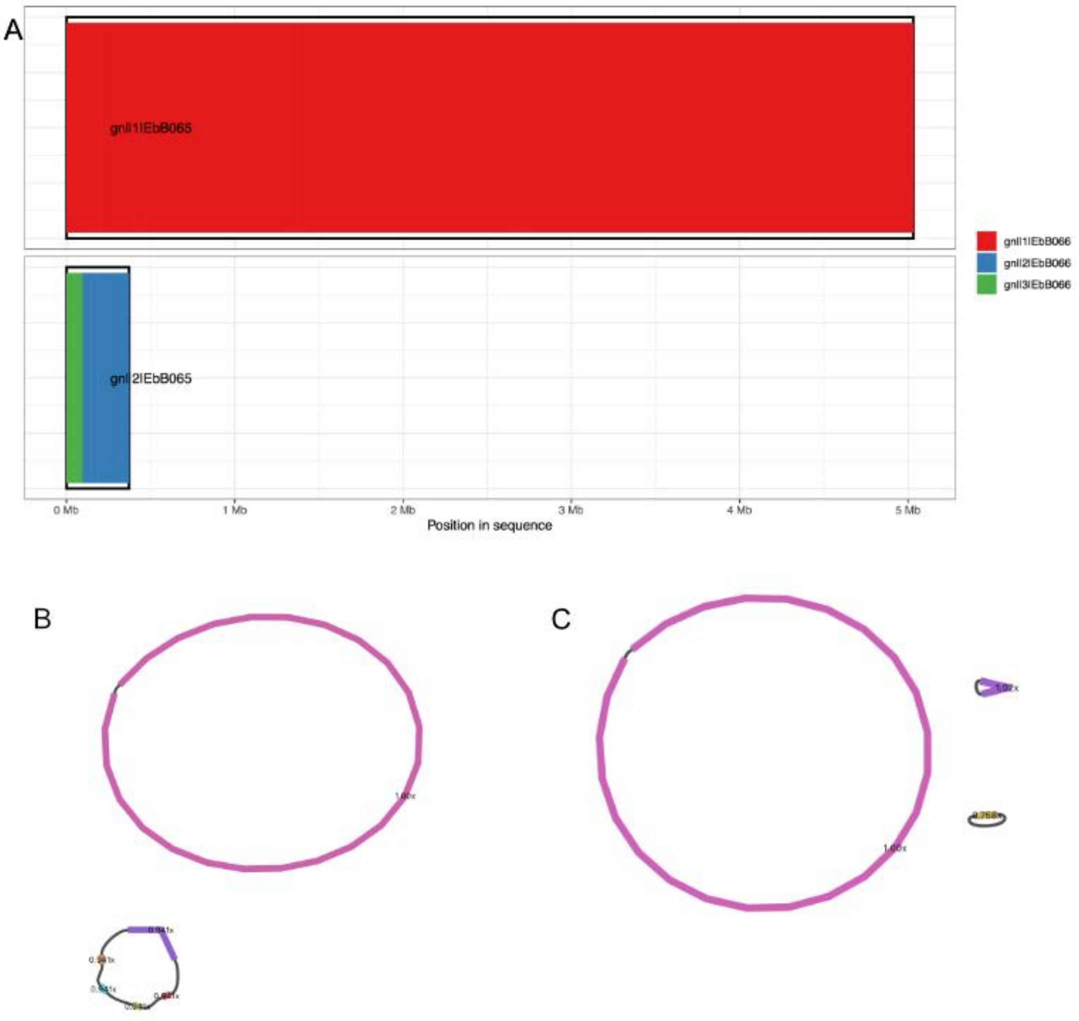

Supplementary Figure 3. A) Alignment between the three contigs assembled in strain EbB066, represented by different coloured blocks as shown in the legend, and the two contigs assembled in strain EbB065. The two non-chromosomal contigs in strain EbB066 are present as one contig in strain EbB065. Bandage visualization of the assembly graph for B) EbB065 and C) EbB066.

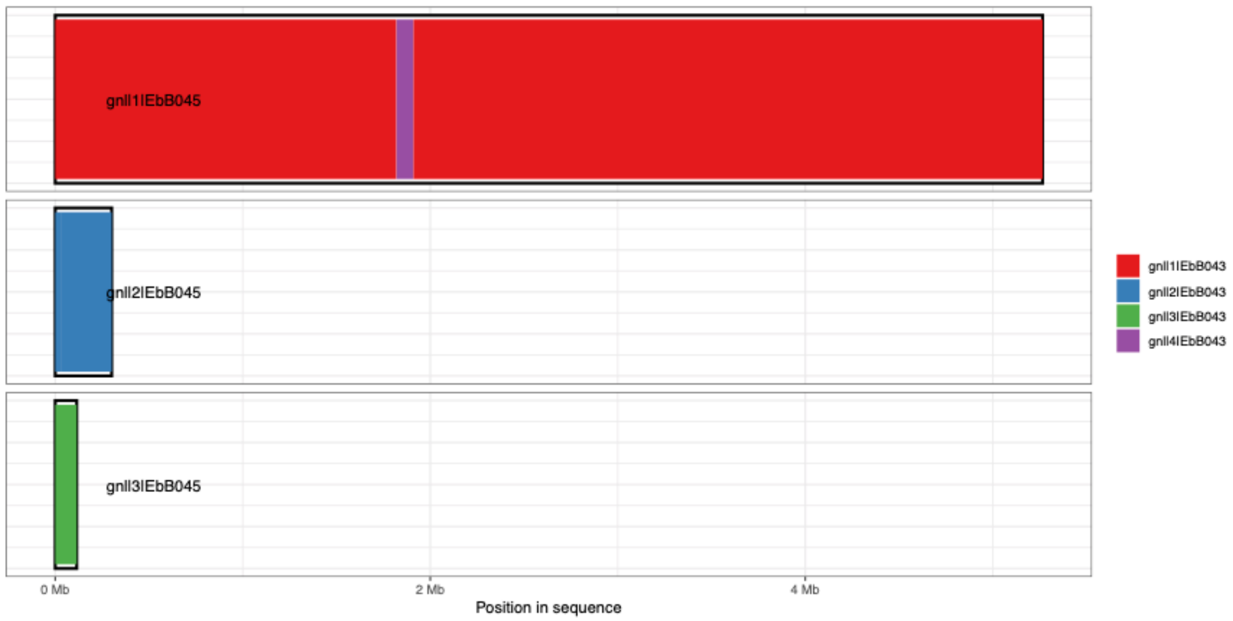

Supplementary Figure 4. A) Alignment between the chromosome and three contigs assembled in strain EbB043, represented by different coloured blocks as shown in the legend, and the chromosome and two contigs assembled in strain EbB045. A non-circular contig assembled in EbB043 is inserted into the chromosome of EbB045.

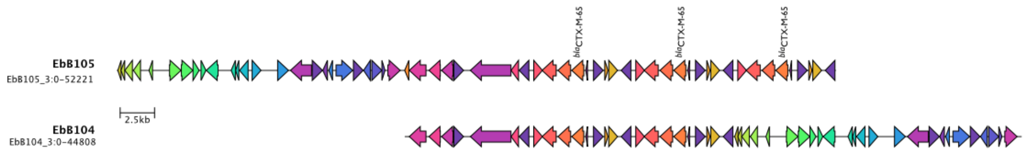

Supplementary Figure 5. Alignment of plasmid region of EbB104 and EbB105 strains showing the tandem duplication of the *bla*$_{CTX-M-65}$ gene (labelled) seen in two copies in EbB104 and three copies in EbB105.

#### Supplementary tables

| Lab ID | Patient ID | Mother or infant? | Organism | Nanopore sequencing | MLST | Transmission pair? |
|---|---|---|---|---|---|---|
| EbB001 | MIC-M-F-0001 | MOTHER | *Escherichia coli* | Yes | 10 | |
| EbB002 | MIC-M-F-0002 | MOTHER | *Escherichia coli* | Yes | 4040 | |
| EbB003 | MIC-I-S-0002 | INFANT | *Klebsiella pneumoniae* | | 667 | |
| EbB004 | MIC-M-F-0003 | MOTHER | *Escherichia coli* | Yes | 131 | |
| EbB005 | MIC-M-F-0003 | MOTHER | *Klebsiella pneumoniae* | | - | |
| EbB006 | MIC-M-F-0004 | MOTHER | *Escherichia coli* | Yes | 2914 | |
| EbB007 | MIC-M-F-0005 | MOTHER | *Escherichia clade-1* | | 7160 | |
| EbB008 | MIC-M-F-0005 | MOTHER | *Escherichia fergusonii* | | 5643 | |
| EbB009 | MIC-M-F-0006 | MOTHER | *Escherichia coli* | | 2852 | |
| EbB010 | MIC-M-F-0006 | MOTHER | *Escherichia coli* | | 48 | |
| EbB012 | MIC-M-F-0006 | MOTHER | *Klebsiella pneumoniae* | Yes | 37 | |
| EbB013 | MIC-I-S-0007 | INFANT | *Escherichia coli* | Yes | 38 | |
| EbB014 | MIC-M-F-0007 | MOTHER | *Escherichia coli* | | 758 | |
| EbB015 | MIC-M-F-0008 | MOTHER | *Escherichia coli* | Yes | 2067 | |
| EbB016 | MIC-M-F-0008 | MOTHER | *Escherichia coli* | | 165 | |
| EbB018 | MIC-I-S-0009 | INFANT | *Escherichia coli* | Yes | 4040 | |
| EbB019 | MIC-M-F-0009 | MOTHER | *Escherichia coli* | Yes | 48 | |
| EbB020 | MIC-M-F-0009 | MOTHER | *Escherichia coli* | | 361 | |
| EbB021 | MIC-M-F-0009 | MOTHER | *Escherichia fergusonii* | | - | |
| EbB022 | MIC-M-F-0009 | MOTHER | *Klebsiella pneumoniae* | | - | |
| EbB023 | MIC-I-S-0010 | INFANT | *Escherichia coli* | Yes | 58 | Yes, with EbB023 |
| EbB024 | MIC-M-F-0010 | MOTHER | *Escherichia coli* | Yes | 58 | Yes, with EbB024 |
| EbB025 | MIC-M-F-0010 | MOTHER | *Klebsiella pneumoniae* | Yes | 20 | |
| EbB026 | MIC-M-F-0012 | MOTHER | *Escherichia coli* | | 414 | |
| EbB027 | MIC-I-S-0012 | INFANT | *Klebsiella pneumoniae* | | 188 | |
| EbB028 | MIC-I-S-0012 | INFANT | *Klebsiella quasipneumoniae subsp. similipneumoniae* | | 1226 | |
| EbB029 | MIC-I-S-0014 | INFANT | *Escherichia coli* | | 405 | |

| | | | | | | |
|---|---|---|---|---|---|---|
| EbB030 | MIC-M-F-0014 | MOTHER | *Escherichia coli* | | 3052 | |
| EbB031 | MIC-I-S-0015 | INFANT | *Escherichia coli* | Yes | 44 | |
| EbB032 | MIC-I-S-0015 | INFANT | *Escherichia coli* | Yes | 5947 | |
| EbB033 | MIC-M-F-0015 | MOTHER | *Escherichia clade-1* | | - | |
| EbB034 | MIC-M-F-0015 | MOTHER | *Escherichia coli* | | 162 | |
| EbB035 | MIC-M-F-0015 | MOTHER | *Klebsiella pneumoniae* | | - | |
| EbB036 | MIC-I-S-0016 | INFANT | *Escherichia coli* | Yes | 38 | Yes, with EbB037 |
| EbB037 | MIC-M-F-0016 | MOTHER | *Escherichia coli* | Yes | 38 | Yes, with EbB036 |
| EbB038 | MIC-M-F-0016 | MOTHER | *Klebsiella pneumoniae* | | 500 | |
| EbB039 | MIC-M-F-0019 | MOTHER | *Escherichia coli* | Yes | 131 | |
| EbB041 | MIC-I-S-0020 | INFANT | *Escherichia coli* | Yes | 44 | |
| EbB042 | MIC-M-F-0020 | MOTHER | *Escherichia coli* | Yes | 38 | |
| EbB043 | MIC-I-S-0020 | INFANT | *Klebsiella pneumoniae* | Yes | 36 | |
| EbB044 | MIC-I-S-0020 | INFANT | *Klebsiella pneumoniae* | | 1322 | |
| EbB045 | MIC-M-F-0020 | MOTHER | *Klebsiella pneumoniae* | Yes | 36 | |
| EbB046 | MIC-M-F-0023 | MOTHER | *Escherichia coli* | | 226 | |
| EbB047 | MIC-M-F-0025 | MOTHER | *Escherichia coli* | | 361 | |
| EbB048 | MIC-M-F-0025 | MOTHER | *Escherichia coli* | | 2083 | |
| EbB049 | MIC-M-F-0026 | MOTHER | *Escherichia coli* | Yes | 457 | |
| EbB050 | MIC-M-F-0026 | MOTHER | *Escherichia coli* | | 410 | |
| EbB051 | MIC-I-S-0027 | INFANT | *Escherichia coli* | Yes | 224 | Yes, with EbB052 |
| EbB052 | MIC-M-F-0027 | MOTHER | *Escherichia coli* | Yes | 224 | Yes, with EbB051 |
| EbB053 | MIC-M-F-0027 | MOTHER | *Klebsiella pneumoniae* | | 17 | |
| EbB054 | MIC-I-S-0028 | INFANT | *Escherichia coli* | | 617 | |
| EbB055 | MIC-M-F-0028 | MOTHER | *Escherichia coli* | | 10 | |
| EbB056 | MIC-M-F-0028 | MOTHER | *Escherichia coli* | | 2705 | |
| EbB057 | MIC-M-F-0028 | MOTHER | *Klebsiella quasipneumoniae subsp. similipneumoniae* | | 334 | |
| EbB058 | MIC-M-F-0032 | MOTHER | *Escherichia coli* | | 226 | |
| EbB060 | MIC-I-S-0033 | INFANT | *Escherichia coli* | | 6778 | |
| EbB061 | MIC-I-S-0033 | INFANT | *Escherichia coli* | | 3075 | |
| EbB062 | MIC-M-F-0033 | MOTHER | *Escherichia coli* | | 515 | |

| | | | | | | |
|---|---|---|---|---|---|---|
| EbB063 | MIC-M-F-0033 | MOTHER | *Escherichia coli* | | 10 | |
| EbB064 | MIC-M-F-0033 | MOTHER | *Klebsiella pneumoniae* | | 76 | |
| EbB065 | MIC-I-S-0034 | INFANT | *Escherichia coli* | Yes | 354 | Yes, with EbB066 |
| EbB066 | MIC-M-F-0034 | MOTHER | *Escherichia coli* | Yes | 354 | Yes, with EbB065 |
| EbB067 | MIC-M-F-0034 | MOTHER | *Escherichia coli* | | 746 | |
| EbB068 | MIC-M-F-0034 | MOTHER | *Klebsiella quasipneumoniae subsp. similipneumoniae* | | - | |
| EbB069 | MIC-M-F-0039 | MOTHER | *Escherichia coli* | | 973 | |
| EbB070 | MIC-I-S-0039 | INFANT | *Klebsiella quasipneumoniae subsp. quasipneumoniae* | | 2823 | |
| EbB071 | MIC-M-F-0039 | MOTHER | *Klebsiella pneumoniae* | | 17 | |
| EbB075 | MIC-M-F-0040 | MOTHER | *Klebsiella pneumoniae* | | 17 | |
| EbB076 | MIC-M-F-0041 | MOTHER | *Escherichia coli* | | 48 | |
| EbB077 | MIC-M-F-0041 | MOTHER | *Escherichia coli* | | 131 | |
| EbB091 | MIC-I-S-0051 | INFANT | *Escherichia coli* | Yes | 38 | |
| EbB092 | MIC-M-F-0051 | MOTHER | *Escherichia coli* | | - | |
| EbB093 | MIC-M-F-0051 | MOTHER | *Escherichia fergusonii* | | - | |
| EbB094 | MIC-M-F-0051 | MOTHER | *Klebsiella pneumoniae* | | 307 | |
| EbB095 | MIC-I-S-0052 | INFANT | *Escherichia coli* | | 68 | |
| EbB096 | MIC-M-F-0052 | MOTHER | *Escherichia coli* | Yes | 1788 | |
| EbB097 | MIC-M-F-0052 | MOTHER | *Escherichia coli* | Yes | 1788 | |
| EbB099 | MIC-M-F-0056 | MOTHER | *Escherichia coli* | | 648 | |
| EbB101 | MIC-M-F-0060 | MOTHER | *Escherichia coli* | | 10 | |
| EbB102 | MIC-I-S-0061 | INFANT | *Escherichia coli* | Yes | 12 | Yes, with EbB103 |
| EbB103 | MIC-M-F-0061 | MOTHER | *Escherichia coli* | Yes | 12 | Yes, with EbB102 |
| EbB104 | MIC-I-S-0061 | INFANT | *Klebsiella pneumoniae* | Yes | 273 | Yes, with EbB105 |
| EbB105 | MIC-M-F-0061 | MOTHER | *Klebsiella pneumoniae* | Yes | 273 | Yes, with EbB104 |
| EbB106 | MIC-I-S-0062 | INFANT | *Escherichia coli* | Yes | 3268 | Yes, with EbB107 |
| EbB107 | MIC-M-F-0062 | MOTHER | *Escherichia coli* | Yes | 3268 | Yes, with EbB106 |
| EbB108 | MIC-M-F-0064 | MOTHER | *Escherichia coli* | | 5846 | |
| EbB109 | MIC-I-S-0065 | INFANT | *Escherichia coli* | | 38 | |
| EbB110 | MIC-M-F-0065 | MOTHER | *Escherichia coli* | Yes | 38 | |
| EbB111 | MIC-I-S-0066 | INFANT | *Escherichia coli* | | 189 | |

| | | | | | |
|---|---|---|---|---|---|
| EbB112 | MIC-I-S-0067 | INFANT | *Escherichia coli* | | 5869 |
| EbB113 | MIC-M-F-0067 | MOTHER | *Escherichia coli* | | 141 |
| EbB114 | MIC-I-S-0067 | INFANT | *Klebsiella pneumoniae* | | 163 |
| EbB115 | MIC-M-F-0067 | MOTHER | *Klebsiella pneumoniae* | | 1035 |
| EbB117 | MIC-M-F-0069 | MOTHER | *Escherichia coli* | | 2689 |
| EbB118 | MIC-M-F-0069 | MOTHER | *Escherichia coli* | | 453 |
| EbB120 | MIC-M-F-0070 | MOTHER | *Escherichia coli* | | 315 |

Supplementary Table 1. Samples sequenced in this study.
